# Safety and immunogenicity of SW-BIC-213, a modified COVID-19 Lipo-Polyplex mRNA vaccine, in Laotian healthy adults aged 18 years and above: a Phase 1/2 trial

**DOI:** 10.1101/2023.09.11.23295344

**Authors:** Yujie Chen, Jingxin Li, Davone Duangdany, Chanthala Phamisith, Bo Yu, Shengke Lan, Lairun Jin, Dawei Lv, Yang Li, Bin Luo, Peng Han, Jinyan Wu, Yuzhu Wang, Congcong Xu, Mingyun Shen, Fanfan Zhao, Peipei Liu, Rongjuan Pei, Haifa Shen, Wuxiang Guan, Hangwen Li, Mayfong Mayxay

## Abstract

**Background:** The mRNA vaccine against SARS-CoV-2 has demonstrated remarkable efficacy in protecting against coronavirus disease 2019 (COVID-19), including providing high protection against severe disease during the emergence of variant waves. In this study, we aimed to investigate the safety and immunogenicity of a 2-dose regimen of the LPP-based mRNA vaccine, SW-BIC-213, in Laos.

**Methods:** For this phase 1/2 clinical trial, we recruited healthy adults aged 18-60 years (phase 1) or ≥18 years (phase 2) from Mahosot Hospital (Vientiane) and Champhone District Hospital (Savannakhet). Participants with SARS-CoV-2 infection, previous COVID-19 vaccination, known allergies to any vaccine component, or pregnancy were excluded. In the phase 1 trial, 41 eligible participants were sequentially assigned to either the 25 μg dose group (25 μg) or the 45 μg dose group (45 μg) in accordance with their enrollment order, with 21 participants in 45 μg dose group and 20 participants in 25 μg dose group. In the phase 2 trial, 480 participants were randomly allocated (2:2:1 ratio) to either the 25 μg dose group, 45 μg dose group, or placebo group.

The primary endpoints for the phase 1 trial were the incidence of local/systemic solicited adverse reactions/events (0-6 days after each vaccination dose), unsolicited adverse events (0-21 days and 0-28 days after the first and second dose of immunization, respectively), and serious adverse events from the first dose of vaccination to 28 days after completing the full course of immunization. In the phase 2 trial, the primary endpoints were the seroconversion rate and geometric mean titer (GMT) of SARS-CoV-2 S-protein specific IgG antibodies and neutralizing antibodies 14 days after the second dose in participants. As for neutralizing antibodies, we detected pseudo-virus neutralizing antibody against wild type (WT), Delta, BA.1 and BA.2. We also detected live viral neutralizing antibody against WT strain 14 days after the second dose. Furthermore, the safety endpoints were also measured during the trial.

This seamless phase 1/2 trial was registered with ClinicalTrials.gov under the identifier NCT05144139.

**Results:** Between December 3, 2021, and March 31, 2022, a total of 41 participants were recruited in the phase 1 trial, while the phase 2 trial enrolled 480 participants from January 20 to July 6, 2022. In the phase 1 trial, a total of 32 subjects (80.0%) reported 103 cases of adverse reactions. All adverse reactions were limited to Grade 1-2. In the phase 2 trial, a total of 479 subjects, 372 subjects (77.7%) reported 929 cases of adverse reactions. All adverse reactions in severity of Grade 3 were manifested as fever (3.4%, 2.1% and 2.9% in 45 μg dose, 25 μg dose and placebo group respectively, only observed in adults), except which all other reactions were limited to Grade 1-2. All adverse reactions noted during the study were tolerable, predominantly transient, and resolved spontaneously. No serious adverse events (SAEs) related to vaccination were observed.

In Phase 2 study, SW-BIC-213 could elicit a high level of seroconversion rate of pseudo-virus neutralizing antibody against WT (100.0% in 25 μg dose group, 99.3% in 45 μg dose group), Delta (99.2% in 25 μg dose group, 98.0% in 45 μg dose group), Omicron BA.1 (84.1% in 25 μg dose group, 84.7% in 45 μg dose group) and Omicron BA.2 (96.0% in 25 μg dose group, 88.8% in 45 μg dose group) at 14 days after the second dose. The pseudo-virus neutralizing antibody titer against WT, Delta, BA.1 and BA.2 was all significant higher (P<0.0001) in both 45 μg dose group (1175.02, 620.62, 72.39 and 172.80) and 25 μg dose group (885.80, 579.40, 47.24 and 101.96) compared with the placebo group (9.67, 10.66, 13.99 and 29.53) at 14 days after the second dose. As for live viral neutralizing antibodies against WT strain, the seroconversion rate could reach more than 94% at 14 days after second dose. The neutralizing antibody titer against WT strain was significantly higher (P<0.0001) in both 45 μg dose group (315.00) and 25 μg dose group (323.18) compared with the placebo group (8.51) at 14 days after second dose.

**Conclusion:** COVID-19 mRNA vaccine SW-BIC-213 manifests a favorable safety profile and is highly immunogenic in eligible subjects aged ≥18 years.

## INTRODUCTION

Since the emergence of COVID-19, it has had a huge impact on the whole world^1^. According to statistics from the World Health Organization (WHO), there has been over 767 million confirmed cases of COVID-19, including 6.9 million deaths as of June 2023^2^. One of the best ways to prevent and slow down the transmission is vaccination^3^.

In response to the COVID-19 outbreak caused by the SARS-CoV-2 virus, numerous of vaccines development programs have been launched. As of March 2023, the World Health Organization’s vaccine tracker reported a total of 382 vaccine candidates under development^4^. These candidates utilize various platforms, including live attenuated vaccines, recombinant protein vaccines, vector vaccines (replication-incompetent vector vaccines, replication-competent vector vaccines, and inactivated virus vector vaccines), DNA vaccines and RNA vaccines^5–10^. Among these platforms, mRNA vaccines have demonstrated the highest protective efficacy^5–7^. Notably, BNT162b2 and mRNA-1273 are representative mRNA vaccines against SARS-CoV-2. The BNT162b2 vaccine, administered as a two-dose regimen (30 μg per dose, with a 21-day interval), has shown a safety profile and 95% efficacy against COVID-19^11^. Similarly, the mRNA-1273 vaccine exhibited 94.1% efficacy in preventing COVID-19 illness, including severe disease^12^. These vaccines have made significant contributions in controlling the COVID-19 pandemic and have demonstrated the feasibility of mRNA vaccine technology.

SW-BIC-213 is an investigational mRNA that encodes a modified full-length spike protein of WT SARS-CoV-2, incorporating mutations such as 2P mutation, furin mutation, and D614G mutation^13^. It is based on Lipo-Polyplex (LPP) platform^14^. In this study, we present the clinical trial data from a Phase 1/2 trial among healthy volunteers in Laos (Lao PDR) to assess the safety and immunogenicity of the LPP-based mRNA vaccine against COVID-19.

## METHODS

### Study design and participants

We conducted a seamless Phase 1/2 study in Laos to evaluate the safety and immunogenicity of the COVID-19 mRNA vaccine. The Phase 1 trial employed an open-label, single-arm, dosage-escalating design and included healthy individuals aged 18-60 years. The Phase 2 trial utilized a randomized, double-blind, placebo-controlled design and enrolled healthy individuals aged 18 years and above.

During the screening visit, participants underwent a medical history review and physical examination to ensure overall good health. Both male and female participants of childbearing age agreed to use contraception throughout the trial, and pregnant individuals were excluded. Exclusion criteria included a history of SARS/MERS or SARS-CoV-2 infection, experiencing common clinical features of COVID-19 within 14 days prior to enrollment, allergies to vaccine components, history of seizures or mental illness, acute or chronic diseases in the acute phase, congenital or acquired immunodeficiency, HIV infection, lymphoma, leukemia, or other autoimmune diseases, receipt of blood products within the past 3 months, receipt of other study drugs within the past 6 months, or receipt of any COVID-19 vaccine. The exclusion of SARS/MERS or SARS-CoV-2 infection was achieved by asking about infection history and detecting IgG level (2019-nCoV IgG/IgM Detection Kit, Vazyme). Prior to participation, all individuals received comprehensive information about the trial procedures and potential risks and provided their written informed consent.

The trial received approval from the Ethics Committee and Food & Drug Department of the Ministry of Health, Lao PDR (7745). The protocol and each amendment underwent review by the Independent Ethics Committee of the Lao People’s Democratic Republic Ministry of Health National Ethics Committee for Health Research. The study adhered to the principles outlined in the Declaration of Helsinki and Good Clinical Practice Guidelines. Safety assessment, including the necessity of study suspension or termination, was conducted by an independent Data Safety Monitoring Board (DSMB).

The trial was registered with the identifier NCT05144139 on ClinicalTrials.gov.

### Randomization and blinding

In the Phase 1 trial (open-label), eligible participants were assigned in a 1:1 ratio to receive either a 25 μg dose or a 45 μg dose of SW-BIC-213 based on their enrollment sequence. The trial was carried out step by step from the 25 μg dose group to the 45 μg dose group, the 45 μg dose group was enrolled only when the preliminary 7-day safety assessment of the 25 μg dose group was favorable. The Phase 2 trial (blinded) included two age cohorts: the adult group (≥18 to ≤60 years old) and the seniors group (>60 years old), with a ratio of 3:1. Within each age group, participants were randomized in a 1:1:1 ratio to receive either a 25 μg dose of SW-BIC-213, a 45 μg dose of SW-BIC-213, or a placebo. The randomization of participants into vaccine groups was performed using an interactive response technology (IRT) system. The blocked randomization list was generated by an independent statistician utilizing the SAS system (version 9.4 or later). The staff responsible for vaccine receiving, storing, dispensing, and preparation, who were unblinded, had no further involvement in the trial and were strictly prohibited from disclosing any information that could compromise blinding. Throughout the trial, all other investigators, participants, laboratory staff, and the sponsor remained blinded.

### Procedures

The SW-BIC-213 vaccine was composed of an mRNA encoding full-length spike glycoprotein of the Wuhan-HU-1 isolate of SARS-CoV-2. This mRNA included artificial mutations, such as the K986P/V987P (pre-fusion structure) mutation, the 682-QSAQ-685 mutation (substitution of Furin cleavage site), and the D614G mutation. This vaccine was formulated with lipo-polyplex and was developed and manufactured by StemiRNA Therapeutics Co., Ltd, in Shanghai, China, in accordance with good manufacturing practice guidelines. The vaccine was supplied as a buffered-liquid solution in vials, with each vial containing 50 μg of the vaccine in 0.5 mL. It was stored at temperatures ranging from -25°C to -15°C until use. The placebo consisted of a commercial, pre-packaged preservative-free saline solution containing 0.9% NaCl.

Each participant received two doses of the SW-BIC-213 vaccine candidate, each dose was 25 μg or 45 μg or placebo. The vaccinations were administered in the upper arm deltoid muscle, with an interval of 21 days between each dose. Following each dose, participants were observed on-site for 30 minutes, during which any adverse reactions were recorded by the investigators. Participants documented their body temperature and any adverse events on daily cards during the first 7 days after each vaccination. Participants reported any adverse events occurring within 7-21 and 7-28 days after the first and second vaccination, respectively, using contact cards. Any serious adverse events or pregnancies that developed from the first vaccination until the end of the study were monitored, and follow-up was ongoing. In Phase 1 only, participants underwent blood and urine sample collection before and 4 days after each dose for hematology and chemistry laboratory tests to assess any toxicity following vaccination. The enrollment of each dosage group in the Phase 2 study was carried out only if the corresponding dosage group in the Phase 1 study demonstrated a favorable safety profile.

Solicited local adverse events at the injection site within 7 days after vaccination included pain, pruritus, induration, swelling, redness. Solicited systemic adverse events within 7 days after vaccination included fever, fatigue, headache, myalgia, chills, arthralgia, nausea, swollen lymph nodes (lymphadenopathy), diarrhea, vomiting and acute allergic reaction. Unsolicited adverse events occurring within 0-21 days and 0-28 days after the first and second dose respectively were collexted. Unsolicited adverse events were adverse events that were not included in the protocol-defined solicited adverse reactions. The severity of adverse events were assessed according to Division of AIDS (DAIDS) Table for Grading the Severity of Adult and Pediatric Adverse Events (Version 2.1)^15^. The severity of abnormality in vital sign and laboratory test were primarily evaluated using the Guidelines for Adverse Event Classification Standards for Clinical Trials of Preventive Vaccines issued by NMPA (National Medical Products Administration) in 2019^16^, and further complemented by Toxicity Grading Scale for Healthy Adult and Adolescent Volunteers Enrolled in Preventive Vaccine Clinical Trials issued by the FDA in 2007^17^. The causal association between adverse events and vaccination was final determined by experienced doctors based on actual situation.

Blood samples were collected from participants before each dose of vaccination and 14, 90, and 180 days after the second dose. These samples were used to evaluate antibody responses, specifically Spike/RBD-specific IgG antibodies and neutralizing antibodies against SARS-CoV-2. The titer of total anti-SARS-CoV-2 spike IgG antibodies were determined using an indirect ELISA assay. ELISA plates were coated with recombinant SARS-CoV-2 spike protein. Plates were washed with PBS-T (phosphate buffered saline (PBS) with 0.05% Tween-20) and blocked with 2% goat serum in PBS-T. Serially diluted standard antibody to SARS-CoV-2 spike protein and heat-inactivated human serum samples at a certain dilution were added into wells in duplicate. The bound antigen–antibody complex was detected using a rabbit anti-human IgG horseradish peroxidase (HRP) conjugate, and color development occurred upon the addition of 3,3’ 5,5’-tetramethylbenzidine (TMB) chromogenic substrate. The absorbance was measured using a microplate reader at 450/630 nm. The titer of IgG in serum samples was quantified using a calibration curve and expressed as RU/mL, which was later converted to WHO International Standard titer (NIBSC CODE: 20/136) as binding antibody units per mL (BAU/mL). Additionally, RBD-specific IgG antibodies were detected using a commercial kit from Vazyme (Nanjing, China) using a similar method. The detection range of binding antibody is 20-800 BAU/ml.

The neutralizing antibody titer was determined by a vesicular stomatitis virus (VSV)-based pseudo-virus neutralization assay using commercial kits (Zhongxingrongchuang biotech, Beijing, China). The SARS-CoV-2 pseudo-viruses express full-length spike proteins (Prototype/ Delta/ Omicron BA.1/ Omicron BA.2) and containing a firefly luciferase reporter gene for quantitative measurements of infection by relative light units (RLU). Briefly, SARS-CoV-2 pseudo-virus was preincubated with three-fold serially diluted heat-inactivated human serum samples for 1 hour at 37 °C. The medium was also mixed with pseudo-virus as virus control. Then, Vero cells were inoculated with the respective mixtures in 96-well plates. After incubation for 24h at 37 °C, the cells were lysed, and luciferase activity was measured using a commercial substrate according to the manufacturer’s protocol (Vazyme, Nanjing, China). Luminescence intensity was measured using a microplate reader. The neutralization titer was defined as the reciprocal serum dilution necessary for 50% inhibition of luciferase activity compared with the virus control wells, calculated using the Reed-Muench method. The cut-off value of WT, Delta, BA.1, BA.2 pseudo-virus neutralizing antibody is 8, 8, 5, 6, respectively.

Neutralizing antibody titers were determined using a micro-neutralization assay conducted in a biosafety level III laboratory. Serum samples were serially diluted and then incubated at 37°C for one hour with 100 TCID50 doses of the live SARS-CoV-2 WT strain. The serum-virus mixture was then added to Vero cells and incubated for four days. Cytopathic effects (CPE) were observed under electron microscopy, and the serum dilution factor that resulted in 50% CPE was recorded as the neutralizing antibody titer. The cut-off value of neutralizing antibody in phase 1 trial is 2. The detection rage of neutralizing antibody is 10-4426 in phase 2 trial.

### Outcomes

In the phase 1 trial, the primary outcome focused on the safety of COVID-19 mRNA vaccine in healthy people aged 18-60 years. The secondary outcome assessed the humoral immunogenicity and long-term safety for six months. In the phase 2 trial, the primary outcome aimed to evaluate the humoral immunogenicity and safety in healthy people aged 18 years and above. The secondary outcome examined the long-term safety and persistence of humoral immunity for six months.

Safety outcomes included the incidence of local/systemic solicited adverse reactions/events (0-6 days after each vaccination dose), unsolicited adverse events (0-21 days and 0-28 days after the first and second dose of immunization, respectively), and serious adverse events from the first dose of vaccination to 28 days after completing the full course of immunization.

Outcomes for immunogenicity included seroconversion rate and GMT of SARS-COV-2 spike protein specific IgG antibody and neutralizing antibody (Pseudo-virus neutralizing antibody targeting WT, Delta, Omicron BA.1 and Omicron BA.2 14 days after the second dose; live viral neutralizing antibody against WT strain) 14 days after the second dose. In addition, RBD specific IgG antibody was investigated in phase 1 trial as an exploratory endpoint.

### Statistical analysis

The safety analysis included all participants who received at least one dose of the investigational vaccine. Immunogenicity analyses were primarily based on the per protocol set (PPS) and supplemented by the full analysis set (FAS) at various time points, as defined in the study protocol. The full analysis set comprised all subjects who received at least one dose of vaccination and had valid pre-vaccination antibody results. Moreover, the Per protocol set referred to participants who were enrolled, following the inclusion and exclusion criteria, completed the vaccination course as protocol defined in the protocol, and had valid pre-vaccination and post-vaccination immunogenicity data. Safety endpoints were presented as percentage of participants experiencing adverse events or serious adverse events during the observation period. To compare the differences in proportions across the groups, Fisher’s exact test was used. Immunological analysis for IgG and neutralizing antibodies included seroconversion rates and GMT at specified time points. We calculated 95% confidence intervals using the Clopper-Pearson method. Chi-square test/Fisher’s exact test was used to compare differences in seroconversion rates among the groups. We performed a t-test after log-transformation to compare differences in GMT between the groups. All statistical analyses were conducted using SAS version 9.4.

In phase 1 trial, the probability to observe a particular adverse event with an incidence of 8% at least once in 20 participants in each dose group was 81.1%. The actual enrolled sample size was 41. For phase 2, the sample size calculation was based on the assumption that the seroconversion rate of vaccine and placebo groups be 80% and 10%, respectively, and the test level would be a unilateral α=0·025. We calculated that, with a sample size of 144 subjects in each vaccine group and 36 subjects in each placebo group for adults and 48 subjects in each vaccine group and 12 subjects in each placebo group for seniors, the difference between the vaccine and placebo could be estimated with at least 99% confidence including the consideration of 33% dropout rate. The actual enrolled sample size was 480.

## RESULTS

### Patient characteristics

Between December 3, 2021, and March 31, 2022, a total of 94 individuals underwent recruitment and screening for Phase 1 trial in Vientiane and Savannakhet, Laos (Fig. 1A). After excluding 53 individuals, 41 eligible participants were assigned to two groups, receiving either the 25 μg dose (n=20) or the 45 μg dose (n=21) of the vaccine. Among them, 40 participants (20 in the 45 μg dose group and 20 in the 25 μg dose group) received their first vaccination, and 39 participants (19 in the 45 μg dose group and 20 in the 25 μg dose group) completed the two-dose immunization. The mean age in the 25 μg dose group was 31.3 years, and in the 45 μg dose group was 30.2 years, with a balanced age distribution between the two vaccination groups (Table 1).

**Figure 1.**
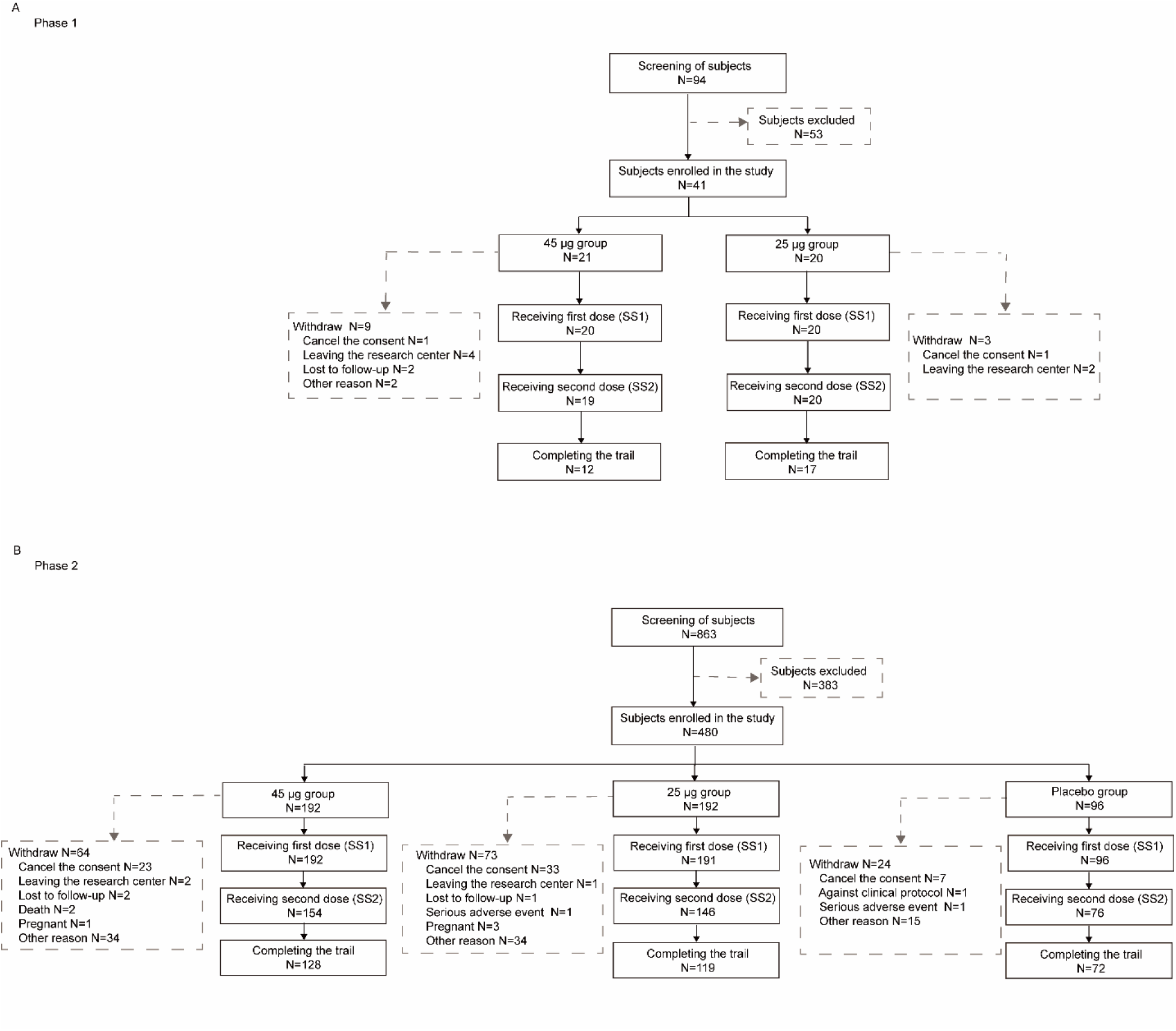
Screening and Randomization of the Participants. The screening and Randomization profile of phase 1 (A) and Phase 2 (B) trial.

**Table 1.**
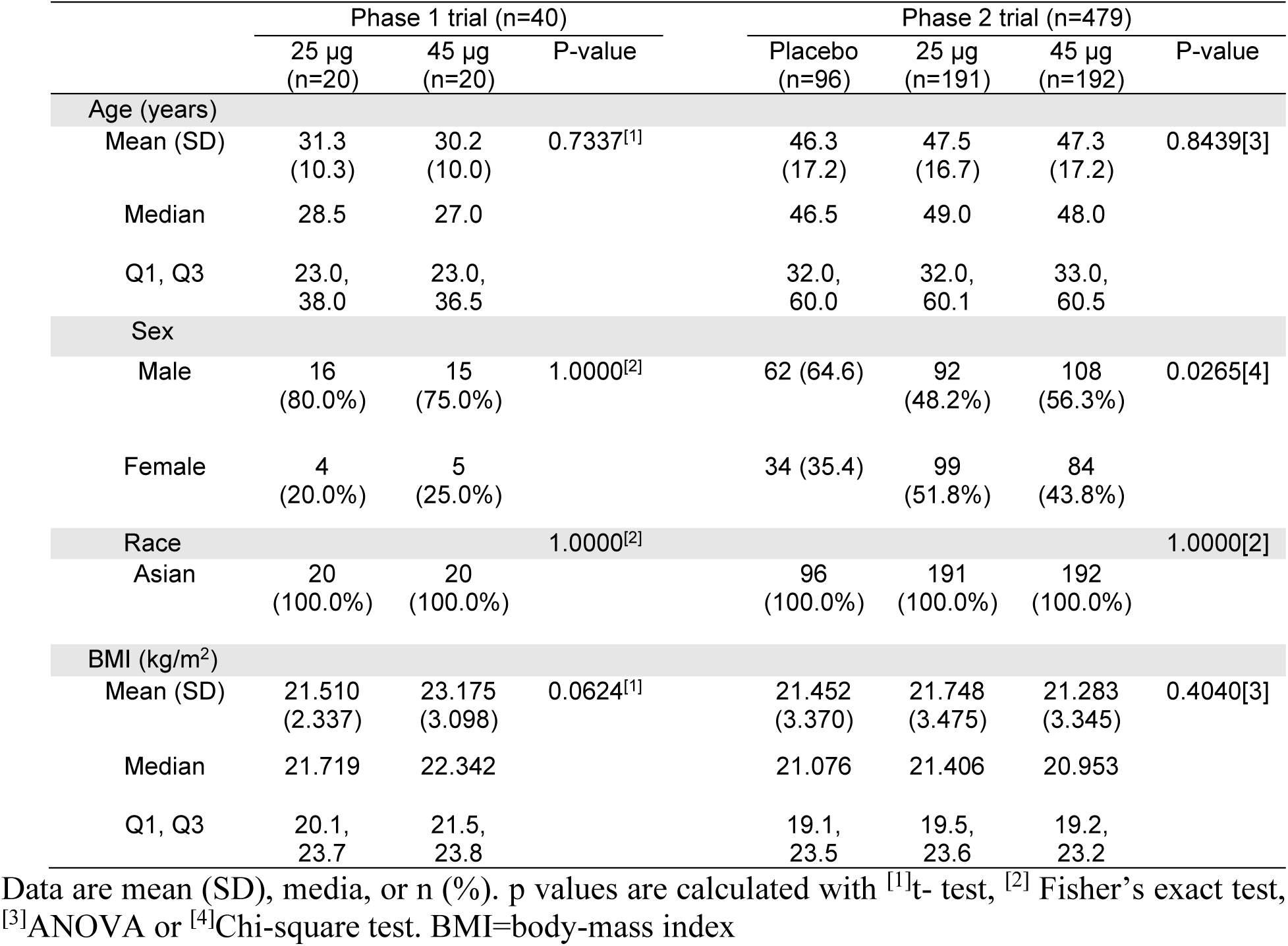
Baseline demographic characteristics of participants in phase 1 and phase 2 trials.

Between January 20, 2022, and July 6, 2022, a total of 863 individuals underwent recruitment and screening for Phase 2 trial in Vientiane and Savannakhet, Laos (Fig. 1B). After excluding 383 individuals, 480 eligible participants were randomly assigned to three groups, receiving either the 25 μg dose (n=192) or the 45 μg dose (n=192) of the vaccine, or placebo (n=96). The mean ages for the placebo group, 25 μg dose group, and 45 μg dose group were 46.3 years, 47.5 years, and 47.3 years, respectively, with a balanced age distribution among the vaccination groups (Table 1). In the Phase 2 study, a total of 479 eligible subjects received their first vaccination on the deltoid muscle, with the investigational vaccine dosage of 25 μg or 45 μg, or placebo. Of them, 376 received the second vaccination using the same immune route and dosage as the first vaccination.

### Safety

The Phase 1 study revealed common adverse reactions, including systemic diseases and various local reactions at the site of administration, such as pain (80.0% in 45 μg dose group and 50.0% in 25 μg dose group), fever (25.0% in 45 μg dose group and 30.0% in 25 μg dose group), fatigue (30.0% in 45 μg dose group and 15.0% in 25 μg dose group), swelling (10.0% in 45 μg dose group and 5.0% in 25 μg dose group), and headache (35.0% in 45 μg dose group and 5.0% in 25 μg dose group). These reactions were limited to grade 1 or 2, with only the occurrence of headache showing a significant difference between the 25 μg dose group and 45 μg dose group (P=0.0436) (Fig. 2A). Nonetheless, all adverse symptoms resolved, mostly spontaneously, and their severity remained limited to Grade 1-2. No grade ≥3 adverse reactions were reported until the data cutoff date.

**Figure 2.**
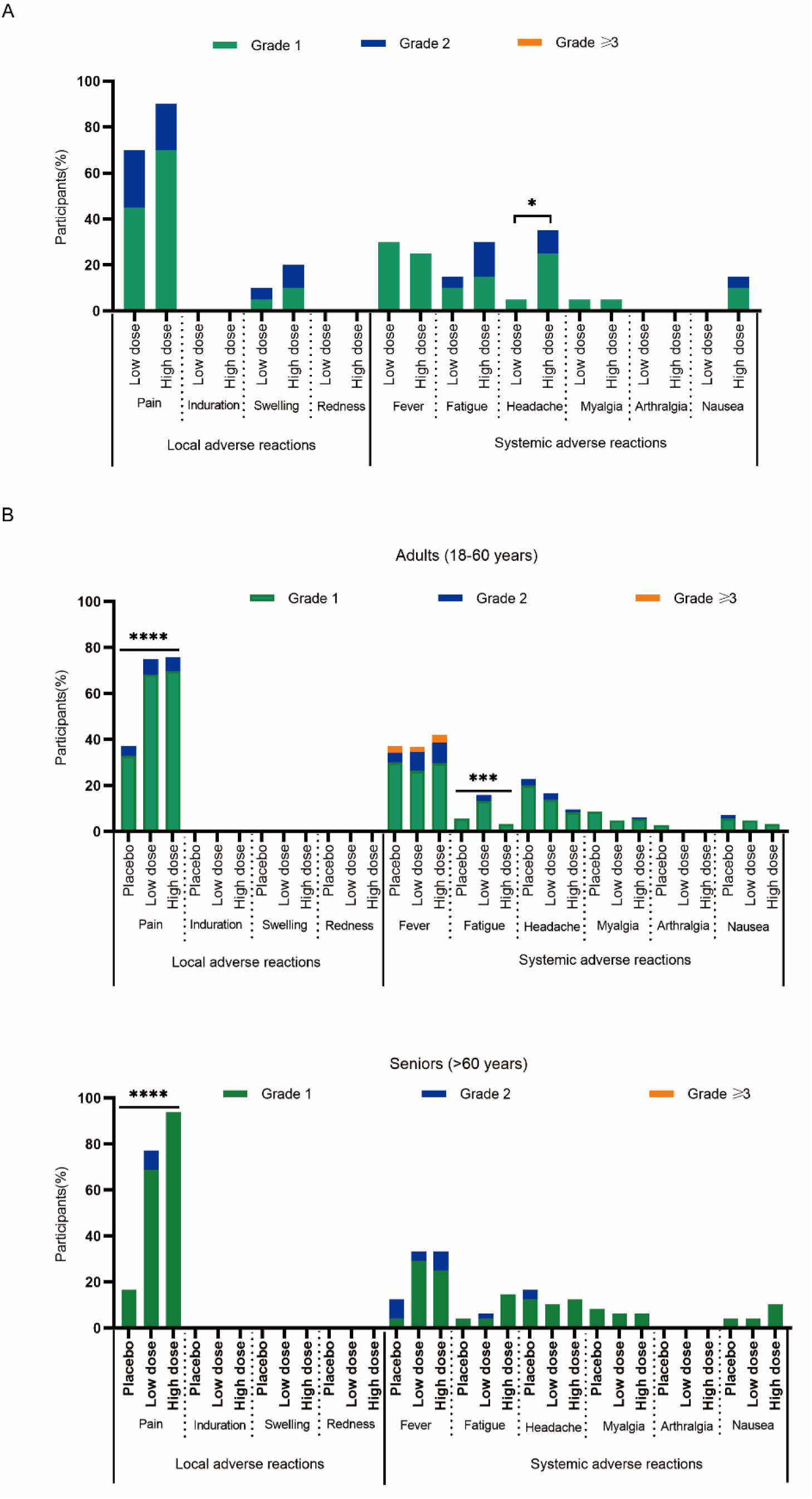
Vaccination-related adverse events and reactions in the phase 1 and phase 2 trials. A. Severity of Solicited Adverse Reactions in phase 1 trial. B. Severity of Solicited Adverse Reactions in phase 2 trial, participants were divided into two age groups, 18-60 years old group (upper panel) and 60 years old above group (lower panel). Severity of the adverse effects were classified according to the NIH guidelines. The Chi-square test was used to assess differences in the incidence of adverse effects among the specified groups. The results of compared analysis among the three groups were expressed as no significant difference (ns, P>0.5), * (p≤0.05), ** (p≤0.01), *** (p≤0.001), **** (p≤0.0001).

Similarly, in the Phase 2 study, common adverse reactions were pain (72.4%, 70.8% and 35.7% for 45 μg dose, 25 μg dose and placebo group, respectively, in adults; 93.8%, 72.9% and 16.7% for 45 μg dose, 25 μg dose and placebo group, respectively, in seniors) at the injection site, fever (37.9%, 36.8% and 35.7% in adults; 33.3%, 33.3% and 12.5% in seniors), fatigue (3.4%, 16.0% and 5.7% in adults; 14.6%, 4.2% and 4.2% in seniors), headache (9.7%, 16.7% and 21.4% in adults; 12.5%, 10.4% and 16.7% in seniors), with primarily grade 1 or 2 intensity. Grade ≥3 adverse reaction was only observed in fever and in adults, the incidence was merely 3.4%, 2.1% and 2.9% in 45 μg dose, 25 μg dose and placebo group respectively. Notably, in the adults group and seniors group, the incidence of pain at the injection site was significantly higher (P<0.0001) in the vaccine group (72.4% and 93.8% for 45 μg dose group, 70.8% and 72.9% for 25 μg dose group) compared to the placebo group (35.7% and 16.7%). Moreover, in the adult group, the incidence of fatigue was significantly higher in the 25 μg dose group compared to the placebo group (P=0.0005) (Fig. 2B). Other adverse effects did not show statistically significant differences between the 45 μg dose group and 25 μg dose group. As with Phase 1, all adverse symptoms resolved, with most of them being transient and self-recovering.

In conclusion, the COVID-19 mRNA vaccine, SW-BIC-213, demonstrated a favorable safety profile and was well tolerated by eligible subjects aged ≥18 years, regardless of the dose and study phase. Local reactions were more prevalent than systemic reactions among all participants. The adverse reactions observed during the trial were generally tolerable, transient, and self-resolving. All grade 3 adverse reactions were characterized as grade 3 fever and were successfully treated.

### Immunogenicity assessments

In the Phase 1 trial, we assessed antibody responses by measuring participants’ serological spike-binding IgG titer using ELISA. Comparing data from each vaccination, the post-vaccination GMT in both groups showed an ascending trend from baseline to 21 days after the first dose, and further increased by 14 days after the second dose (Fig. 3A). In 45 μg dose group, GMT increased from 12.82 BAU/ml at baseline, through 306.70 after receiving the first dose, to 1853.40 BAU/ml after receiving the second dose. In 25 μg dose group, GMT increased from 11.46 BAU/ml at baseline, through 275.71 BAU/ml after receiving the first dose, to 924.17 BAU/ml after receiving the second dose (Fig. 3A). Although the GMT of the 45 μg dose group was higher than that of the 25 μg dose group 14 days after the second dose, no statistically significant difference was observed (P=0.0531) (Fig. 3A). The seroconversion rate of S-protein antibody after the first vaccination was 100.0% in both the 45 μg dose group and 25 μg dose group, while after the second vaccination, it was 100.0% in the 45 μg dose group and 94.7% in the 25 μg dose group (Fig. 3B). No significant differences were found between the groups (P = 1.0000). Additionally, both the 45 μg dose group and 25 μg dose group exhibited 100.0% seroconversion rates for S-protein specific antibody at both 90 days and 180 days after full immunization (Fig. 3B).

**Figure 3.**
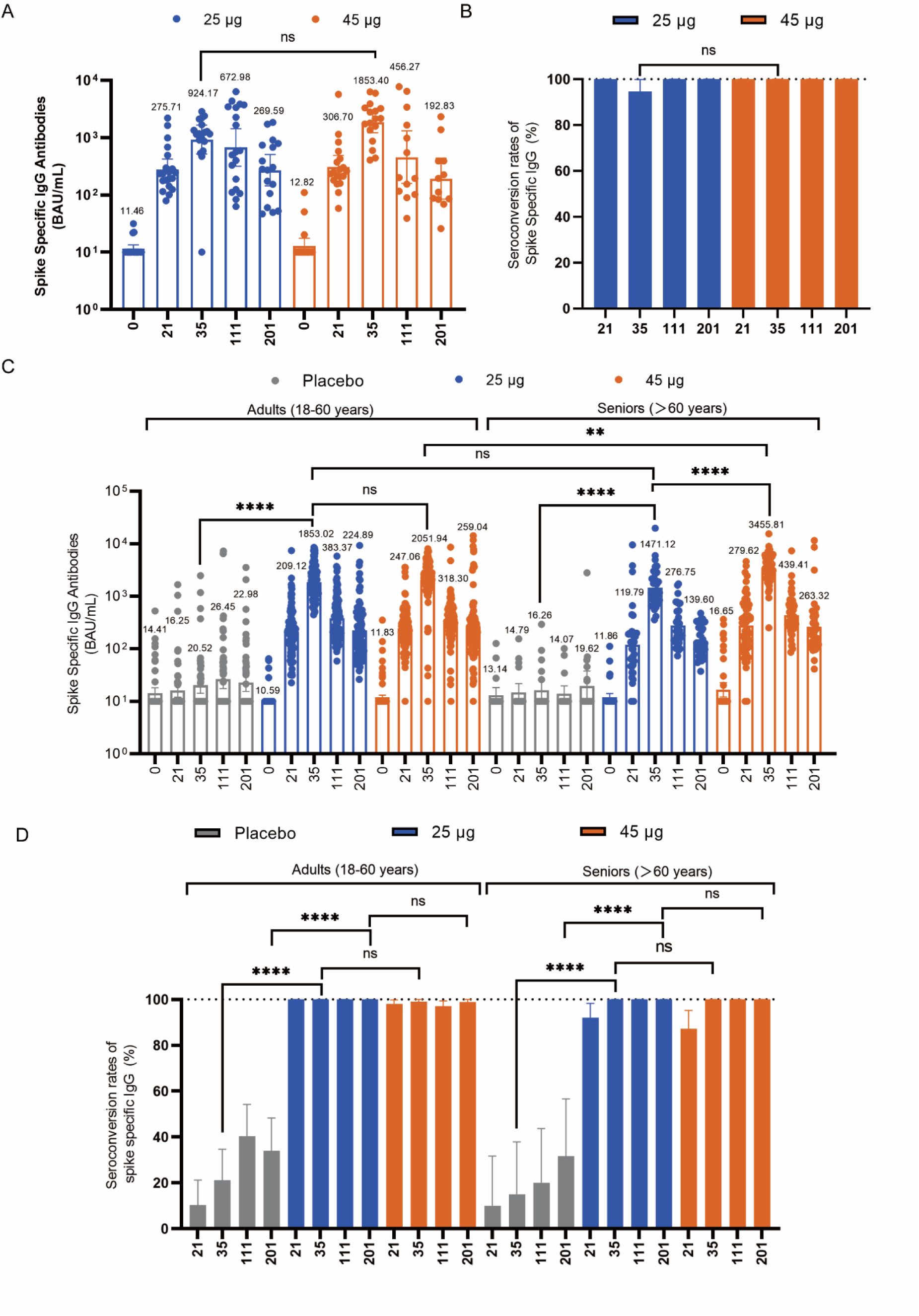
SW-BIC-213 induced Spike protein binding antibodies in phase 1 and phase 2 trials. A. Geometric mean SARS-CoV-2 spike protein binding antibodies titers determined by ELISA at the indicated time points (day) post vaccination in phase 1 trial. B. Seroconversion rates at the indicated time points (day) post vaccination in phase 1 trial. C. Geometric mean SARS-CoV-2 spike protein binding antibodies titers determined by ELISA at the indicated time points (day) post vaccination in phase 2 trial. D. Seroconversion rates at the indicated time points (day) post vaccination in phase 2 trial. Seroconversion is defined as Titer above or equal 20 in baseline-seronegative subjects or at least 4-fold rise in baseline-seropositive subjects. Statistical analysis was conducted with t test to compare the log transformed GMTs of the indicated group. Chi-square test was used to analyze the difference of seroconversion rate of the indicated group. The results of compared analysis were expressed as no significant difference (ns, P>0.5), * (p≤0.05), ** (p≤0.01), *** (p≤0.001), **** (p≤0.0001).

Given the importance of the RBD domain of the S protein for viral entry, antibodies targeting this domain of SARS-CoV-2 were expected to be neutralizing and potentially protective. Therefore, we also measured RBD-binding IgG antibodies using ELISA. Similar to the spike-binding IgG antibodies, GMT post-vaccination in both groups showed an ascending trend from baseline through 21 days after the first dose to 14 days after the second dose (Supplementary Fig. 1A). In 45 μg dose group, GMT increased from 12.34 BAU/ml at baseline, through 138.06 BAU/ml after receiving the first dose, to 1988.90 BAU/ml after receiving the second dose. In 25 μg dose group, GMT increased from 10.46 BAU/ml, through 154.79 BAU/ml after receiving the first dose, to 1127.61 BAU/ml after receiving the second dose (Supplementary Fig. 1A). After completing full immunization, the GMT in the 45 μg dose group was higher than that in the 25 μg dose group, but no significant differences were observed between the groups (P = 1.0000). The seroconversion rate of S-protein antibody after the first vaccination was 94.7% in the 45 μg dose group and 100.0% in the 25 μg dose group, while after the second vaccination, it was 100.0% in the 45 μg dose group and 94.7% in the 25 μg dose group (Supplementary Fig. 1B). Seroconversion rates of RBD combining antibody in both the 45 μg dose group and 25 μg dose group remained at 100.0% at both 90 days and 180 days after full immunization (Supplementary Fig. 1B), with no significant differences between the groups (P = 1.0000).

In the Phase 2 trial, adults and seniors in the 45 μg dose group and 25 μg dose group showed an increasing trend in GMT of S-protein specific antibody from baseline through 21 days after the first dose to 14 days after the second dose (Fig. 3C). Among younger subjects, the GMT of S-protein specific antibody was significantly higher in the experimental group compared to the placebo group after each dose, especially detected at 14 days after the second dose (2051.94 BAU/mL, 1853.02 BAU/mL and 20.52 BAU/mL in 45 μg dose group, 25 μg dose group and placebo group, respectively, P<0.0001) (Fig. 3C). However, no significant difference (P=0.1307) was observed between the 45 μg dose group and 25 μg dose group 14 days after the second dose (Fig. 3C). Among seniors group, the GMT of S-protein specific antibody was significantly higher in the experimental group compared to the placebo group after each dose, detected at 14 days after the second dose (3455.81 BAU/mL, 1471.12 BAU/mL and 16.26 BAU/mL in 45 μg dose group, 25 μg dose group and placebo group, respectively, P<0.0001), with the 45 μg dose group demonstrating notably higher GMT compared to the 25 μg dose group (P<0.0001) (Fig. 3C). Among younger subjects who received the experimental vaccine, the first dose of vaccination (98.1% for 45 μg dose group, 100% for 25 μg dose group) induced a comparable seroconversion rate to that after the second dose (99.0% in 45 μg dose group, 100% in 25 μg dose group) (Fig. 3D). The seroconversion rate induced by the experimental vaccine was significantly higher than that elicited by the placebo (19.4%) at 14 days after the second dose (P<0.0001), but there was no significant difference (P=1.0000) between the 45 μg dose group (99.3%) and 25 μg dose group (100.0%). When comparing between age cohorts, the seroconversion rates after full immunization in both cohorts were generally balanced, with both reaching higher than or equal to 99.0%. At 90 days and 180 days after full immunization, the seroconversion rates of S-protein specific antibody in both the 45 μg dose group and 25 μg dose group in each age cohort remained at a level of ≥97% (Fig. 3D).

The serum samples of participants were also assessed for pseudo-virus neutralizing titers against SARS-CoV-2 prototype strain and three variants of concern (VOC) strains (Delta, Omicron BA.1, and Omicron BA.2). In the Phase 1 trial, the GMT of pseudo-virus neutralizing antibody against WT and Delta strain reached 990.50 and 642.08 in 45 μg dose group at 14 days after the second dose, 507.17 and 352.98 in 25 μg dose group respectively (Fig. 4A, B). As for BA.1 and BA.2, the GMT at 14 days after the second dose was relatively lower, which was 35.27 and 55.88 in 45 μg dose group, 25.12 and 36.27 in 25 μg dose group (Fig. 4C, D). After receiving two doses of vaccination, the GMTs for all strains in the 45 μg dose group were consistently higher than that in the 25 μg dose group, but there was no significant difference (P>0.05). At 14 days after full immunization, in the 45 μg dose group, the seroconversion rate of neutralizing antibodies against WT, Delta, BA.1, and BA.2 were 100.0%, 100.0%, 82.4%, and 82.4%, respectively (Supplementary Fig. 2A, B, C, D). In the 25 μg dose group, these rates were 94.7%, 94.7%, 84.2%, and 47.4%, respectively (Supplementary Fig. 2 A, B, C, D). Except for BA.2 (P=0.0291), seroconversion rates for other strains were balanced between groups (P>0.05) (Supplementary Fig. 2A, B, C, D). The GMT of neutralizing antibodies against each strain in each group declined from 90 days to 180 days after the second dose, except for neutralizing antibody against Omicron BA.1 in the 25 μg dose group, which slightly rose during this period (Fig. 4A, B, C, D). This trend was in line with the seroconversion rate analysis, suggesting that it might be attributed to the activation of immune memory cells by exposure to the pathogen or the hybrid immunity induced by infection. In the Phase 2 trial, the vaccine group showed a significant increase in pseudo-virus neutralizing antibodies against WT, Delta, BA.1, and BA.2 compared to the placebo group (Fig. 4E, F, G, H). At 14 days after the second dose, the GMTs of antibodies against each strain in the 45 μg dose group (1078.75, 568.87, 70.30 and 146.37 for WT, Delta, BA.1 and BA.2, respectively, in adults; 1417.10, 751.08, 77.20 and 248.63 for WT, Delta, BA.1 and BA.2, respectively, in seniors) were generally higher or equal to that in the 25 μg dose group (825.85, 576.59, 45.51 and 91.11 for WT, Delta, BA.1 and BA.2, respectively, in adults; 1088.26, 587.73, 52.71 and 141.88 for WT, Delta, BA.1 and BA.2, respectively, in seniors). Particularly for neutralizing antibody against Omicron BA.1 (P=0.0262) and Omicron BA.2 (P=0.0399) in the younger cohort, the 45 μg dose group displayed a significantly higher response (Fig. 4E, F, G, H). The vaccine could elicit relatively higher or generally balanced GMT in seniors (Fig. 4E, F, G, H), which demonstrated that this vaccine was efficient in seniors group. The seroconversion rate of antibodies 14 days after the second dose against each strain were balanced without significant distinction among senior subjects, between the 45 μg dose group (100.0%, 97.9%, 74.5% and 85.1 for WT, Delta, BA.1 and BA.2, respectively) and 25 μg dose group (100.0%, 100.0%, 81.3% and 93.8 for WT, Delta, BA.1 and BA.2, respectively) (Supplementary Fig. 2 E, F, G, H). However, among the younger cohort, the seroconversion rate of BA.2 antibodies was remarkably higher in the 25 μg dose group than that in the 45 μg dose group (96.8% vs. 89.3%) (Supplementary Fig. 2H), and the seroconversion rate of WT (100.0% vs 99.0%), Dleta (98.9% vs 98.1%), BA.1 (85.1.0% vs 89.3%) was generally balanced (Supplementary Fig. 2 E, F, G). There was no obvious decline in seroconversion rates of each strain from 90 days to 180 days after full immunization for both age cohorts in either the 45 μg dose or 25 μg dose group. The slight increase in the seroconversion rate at certain time points might be attributed to changes in the number of participants or infection during the epidemic.

**Figure 4.**
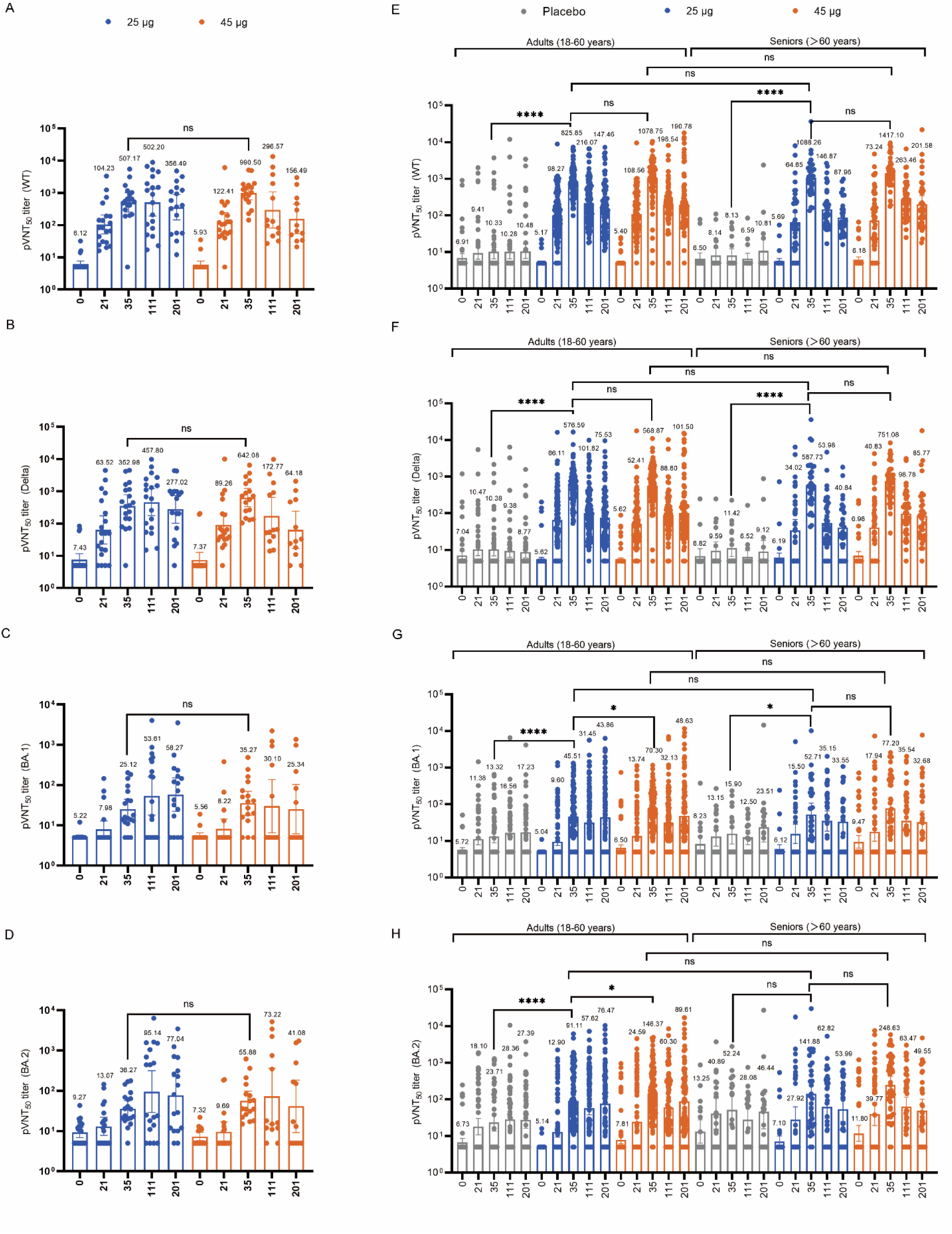
SW-BIC-213 induced pseudo-virus neutralizing antibodies in phase 1 and phase 2 trials. A. GMT of SARS-CoV-2 prototype strain pseudo-virus neutralizing antibody was determined at the indicated time points (day) in phase 1 trial. B. GMT of SARS-CoV-2 delta strain pseudo-virus neutralizing antibody was determined at the indicated time points(day) in phase 1 trial. C. GMT of SARS-CoV-2 omicron BA.1 strain pseudo-virus neutralizing antibody was determined at the indicated time points (day) in phase 1 trial. D. GMT of SARS-CoV-2 omicron BA.2 strain pseudo-virus neutralizing antibody was determined at the indicated time points (day) in phase 1 trial. E. GMT of SARS-CoV-2 prototype strain pseudo-virus neutralizing antibody was determined at the indicated time points (day) in phase 2 trial. F. GMT of SARS-CoV-2 delta strain pseudo-virus neutralizing antibody was determined at the indicated time points(day) in phase 2 trial. G. GMT of SARS-CoV-2 omicron BA.1 strain pseudo-virus neutralizing antibody was determined at the indicated time points (day) in phase 2 trial. H. GMT of SARS-CoV-2 omicron BA.2 strain pseudo-virus neutralizing antibody was determined at the indicated time points (day) in phase 2 trial. Statistical analysis was conducted with t test to compare the log transformed GMTs of the indicated group. The results of compared analysis were expressed as no significant difference (ns, P>0.5), * (p≤0.05), ** (p≤0.01), *** (p≤0.001), **** (p≤0.0001).

In the phase 1 trial, at 14 days after the second dose, the GMT of WT neutralizing antibody was 302.57 in the 45 μg dose group and 161.34 in the 25 μg dose group (Fig. 5A). The seroconversion rates of WT neutralizing antibody were 94.4% in the 45 μg dose group and 100.0% in the 25 μg dose group at 14 days after full immunization (Fig. 5B). There was no significant difference between the 25 μg dose group and the 45 μg dose group (P=0.4737). In the phase 2 trial, when comparing between the adult cohort and senior cohort, the GMT of neutralizing antibody against WT 14 days after full immunization were 270.81 vs. 438.68 in the 45 μg dose group, 352.83 vs. 249.71 in the 25 μg dose group, and 9.44 vs. 6.51 in the placebo group, respectively (Fig. 5C). In all age groups, neutralizing antibody levels were significantly higher in both the 45 μg dose group and 25 μg dose group compared with the placebo group (P<0.0001). Among adults, there was no significant difference between the 45 μg dose group and 25 μg dose group (P=0.0809). However, in the senior group, the neutralizing antibody titer in the 45 μg dose group was significantly higher than that in the 25 μg dose group (P=0.0264). At 14 days post second dose, the seroconversion rates among adult subjects were similar to that among senior subjects. The seroconversion rates were 97.1% vs. 97.9% in the 45 μg dose group, 97.9% vs. 96.9% in the 25 μg dose group, and 21.2% vs. 10.0% in the placebo group (Fig. 5D). Among the age groups, there was a significant difference in the seroconversion rates between the vaccine group and the placebo group (P<0.0001), while there was no significant difference in the seroconversion rates between the 45 μg dose group and the 25 μg dose group (P=0.8442).

**Figure 5.**
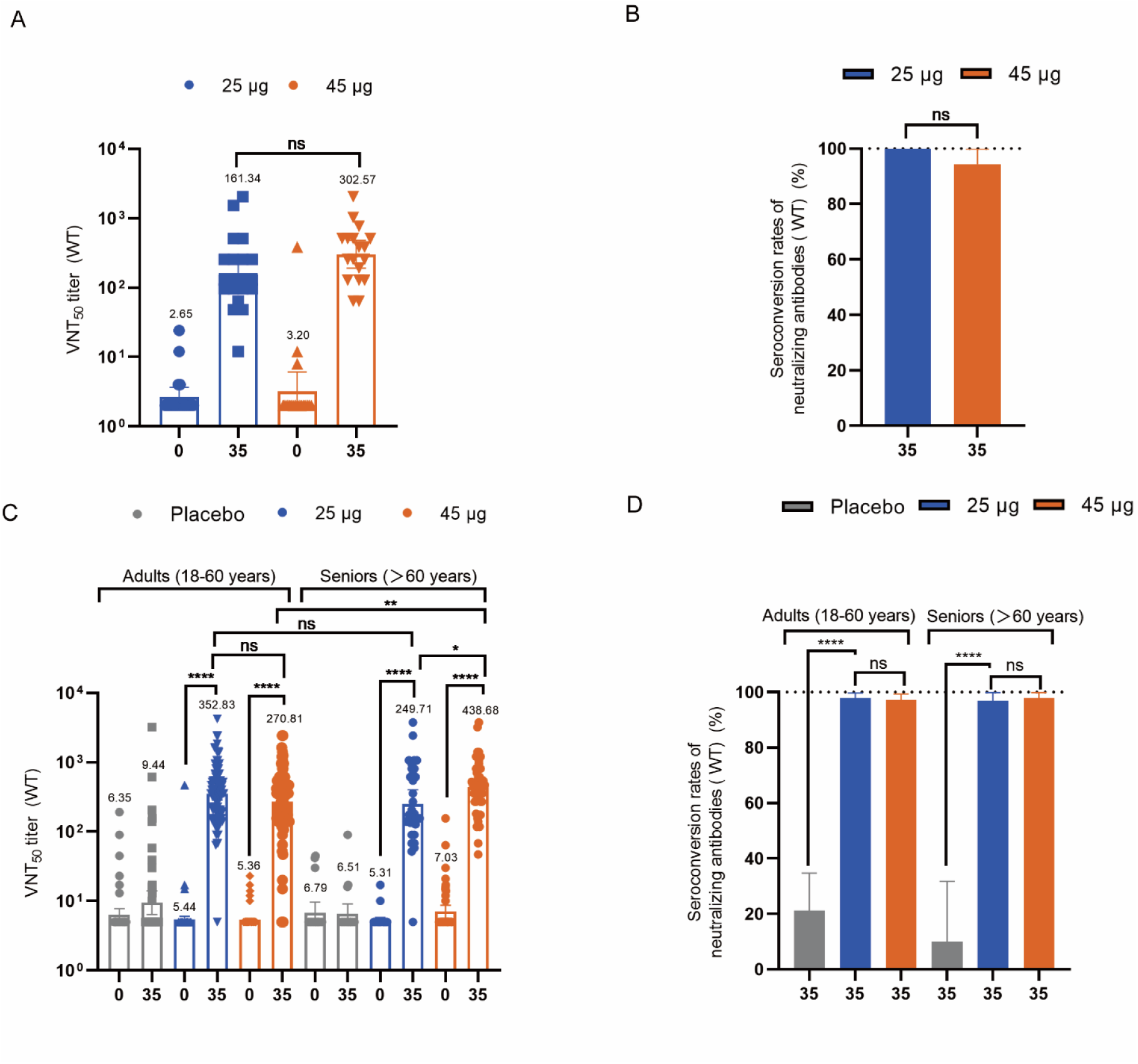
SW-BIC-213 induced neutralizing antibodies in phase 1 and phase 2 trials. A. GMT of SARS-CoV-2 prototype virus neutralizing antibody determined by microneutralization assay in phase 1 trial. B. The seroconversion rate of SARS-CoV-2 prototype virus neutralizing antibody in phase 1 trial. C. GMT of SARS-CoV-2 prototype virus neutralizing antibody determined by microneutralization assay in phase 2 trial. D. The seroconversion rate of SARS-CoV-2 prototype virus neutralizing antibody in phase 2 trial. Seroconversion is defined as titers above or equal 4 in baseline-seronegative subjects or at least 4-fold rise in baseline-seropositive subjects in phase 1 trial. Seroconversion is defined as titers above or equal 10 in baseline-seronegative subjects or at least 4-fold rise in baseline-seropositive subjects in phase 2 trial (the diversity in seroconversion definition from that of phase 2 is attributed to the difference in detection institution and detection standard). Statistical analysis was conducted with t test to compare the log transformed GMTs of the indicated group. Chi-square test was used to analyze the difference of seroconversion rate of the indicated group. The results of compared analysis were expressed as no significant difference (ns, P>0.5), * (p≤0.05), ** (p≤0.01), *** (p≤0.001), **** (p≤0.0001).

## Discussion

In both phase 1 and phase 2 trials, we observed that two-dose SW-BIC-213 with the 25 or 45 μg doses was well tolerated, with all adverse reactions mainly being transient and could resolved spontaneously. Notably, no of vaccination-related SAE or adverse events of special interest, like ADE/VED, was noticed since the first dose. Adverse reactions in both phase 1 and phase 2 were primarily manifested as local solicited reactions, with the most common symptoms being injection-site pain. Importantly, the frequency of solicited systemic adverse reactions events between the vaccine and placebo groups was similar in phase 2. Only a few adults in both the vaccine and placebo groups experienced grade ≥3 adverse reactions in phase 2, notably fever. Other reactions were generally limited to Grade 1-2. Overall, SW-BIC-213 showed favorable safe profile with generally mild or moderate reactogenicity, comparable to two well-known mRNA vaccine, mRNA-1273^18,19^ and BNT162b2^20,21^.

Humoral responses have been considered as immune correlates of protection against SARS-CoV-2^22^. SW-BIC-213 exhibited high immunogenicity and induced robust humoral responses rapidly. Specifically, for S-protein IgG antibody, one dose of vaccination could induce comparable seroconversion rate to that achieved after full immunization, which was roughly equal between 45 μg dose and 25 μg dose group. In contrast, the seroconversion rate of pseudo-virus neutralizing antibody showed an increasing tendency with the progression of the vaccination regimen, reaching higher levels after completing full immunization, regardless of dosage. As for GMT analysis, both IgG and neutralizing antibodies were induced in a dose-dependent manner. Furthermore, as new variants continue to emerge and cause devastation, the neutralizing antibodies against multiple VOC strains were also evaluated. Our candidate was similar potent against Delta strain compared to WT strain with both high seroconversion rate (>97%) and GMT. Meanwhile, in line with previous results^23–27^, a remarkable reduction of neutralizing antibodies was found against the Omicron variants, though the seroconversion rate was high to approximate 80% and 96% for Omicron BA.1 and BA.2 respectively at 14 days post full vaccination. For participants aged 60 years and older, especially with pre-existing respiratory or cardiovascular disease, COVID-19 presents a remarkably high risk of severe disease and death^28^. Notably, in this study, the magnitude of humoral immunogenicity in the group aged above 60 years was comparable to that of in the group aged 18–60 years in the context of similar safety profile.

The long-term tolerability profile of SW-BIC-213 and persistence of the elicited immune responses were also investigated according to the study protocol. Indicated with phase 1 and phase 2 trial result, seroconversion rate of WT neutralizing antibody, Delta neutralizing antibody and S-protein specific antibody could remain at a relatively high level when coming to 90 days and 180 days after full immunization. Situation that seroconversion rate of Omicron BA.1 and Omicron BA.2 neutralizing antibody slightly increased from 90 days to 180 days after full immunization and situation that seroconversion rate of placebo group could reach 50% suggests that there might have been exposure to pathogens or infections during the course of trial and leading to increased antibody levels in individual subjects. Since there was large epidemic in Laos during the trial, this was likely to occur across the groups and did not affect the key conclusion of the comparison between groups. As SARS-CoV-2 continues to evolve and vaccination induced protective immunity declines, the world has been through many waves of COVID-19 pandemic, and many people have experienced breakthrough infections. Those who have been both vaccinated and infected with SARS-CoV-2 can produce much stronger immune response compared to those who have only been vaccinated or infected. Several studies suggest that hybrid immunity confers more effective cross-variant neutralization and more durable protection against new infections^29–31^. The investigational vaccine demonstrated the ability to induce a substantial humoral immune response, characterized by an outstanding seroconversion rate and significant increases in neutralizing titer and S-protein specific antibody from baseline.

This study also had several limitations. Firstly, data interpretation was based on a relatively small sample size and more data from ongoing phase 3 trials will provide further data to comprehensively evaluate the safety and efficacy of SW-BIC-213 as a booster shot in the heterologous boost regimen. Secondly, the study populations were not ethnically diverse, with all participants were of Asian descent. Thirdly, the neutralizing potency against currently circulating VOCs Omicron BA.4/5 was not evaluated in the present due to the constantly shifting of variants of concern, which will be addressed in the ongoing phase 3 trials.

In conclusion, COVID-19 mRNA vaccine SW-BIC-213 manifests a favorable safety profile and was highly immunogenic in all eligible subjects aged ≥18 years of both phase 1 and phase 2 trials, suggesting potential for further investigation as a means of controlling and preventing COVID-19. However, considering that COVID-19 vaccines have been widely used worldwide as primary series, the majority of targeted population may already have some extent of protection, thus a specific dosage of 25 μg would be set for the booster study. Moreover, with the rising number of breakthrough infections of SARS-CoV-2 in previously immunized individuals, there is increasing concern for the need of a booster vaccine dose to combat waning antibody levels and new variants. As a result, an ongoing phase 3 study in Laos is further evaluating SW-BIC-213 as a booster vaccination against the Omicron variant and other emerging VOC in healthy individuals aged 18 years and above, who have already completed the 2-dose series with COVID-19 inactivated vaccines.

## Data Availability

All data produced in the present study are available upon reasonable request to the authors
All data produced in the present work are contained in the manuscript
All data produced are available online at

## Author contributions

M Mayxay is the principal investigator of this trial. D D, C P, B Yu, J-Y Wu and Y-Z Wang participated in designing the trial and study protocol. R-J Pei and P-P Liu contributed to laboratory testing. P Han and M-Y Shen contributed to develop and manufacture SW-BIC-213. Y-J Chen contributed to manuscript writing. S-K Lan, Y Li and B Luo participated in the site work, including the recruitment, participants’ visits and data collection. J-X Li, H-F Shen, W-X Guan and H-W Li contributed to the critical review and revising of the manuscript. D-W Lv, L-R Jin, F-F Zhao and C-C Xu contributed to verify the data. All authors reviewed and approved the final version of the manuscript.

---

**Extended Data Figure 1.**
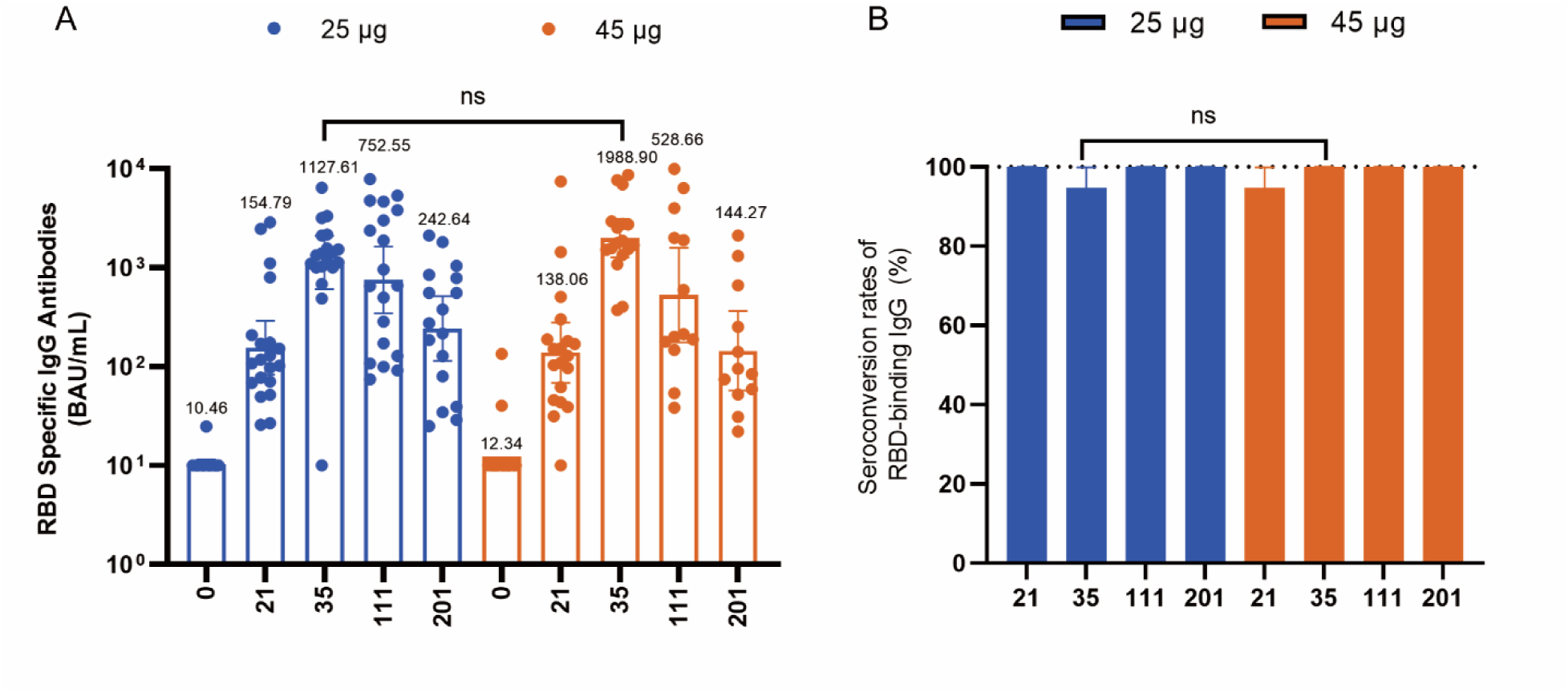
SW-BIC-213 induced RBD binding antibodies in phase 1 trial. A. SARS-CoV-2 RBD protein binding antibody titer determined by ELISA. B. The seroconversion rate of SARS-CoV-2 RBD protein binding antibody. Seroconversion is defined as Titer above or equal 20 in baseline-seronegative subjects or at least 4-fold rise in baseline-seropositive subjects. Statistical analysis was conducted with t test to compare the log transformed GMTs of the indicated group. Chi-square test was used to analyze the difference of seroconversion rate of the indicated group. The results of compared analysis were expressed as no significant difference (ns, P>0.5), * (p≤0.05), ** (p≤0.01), *** (p≤0.001), **** (p≤0.0001).

**Extended Data Figure 2.**
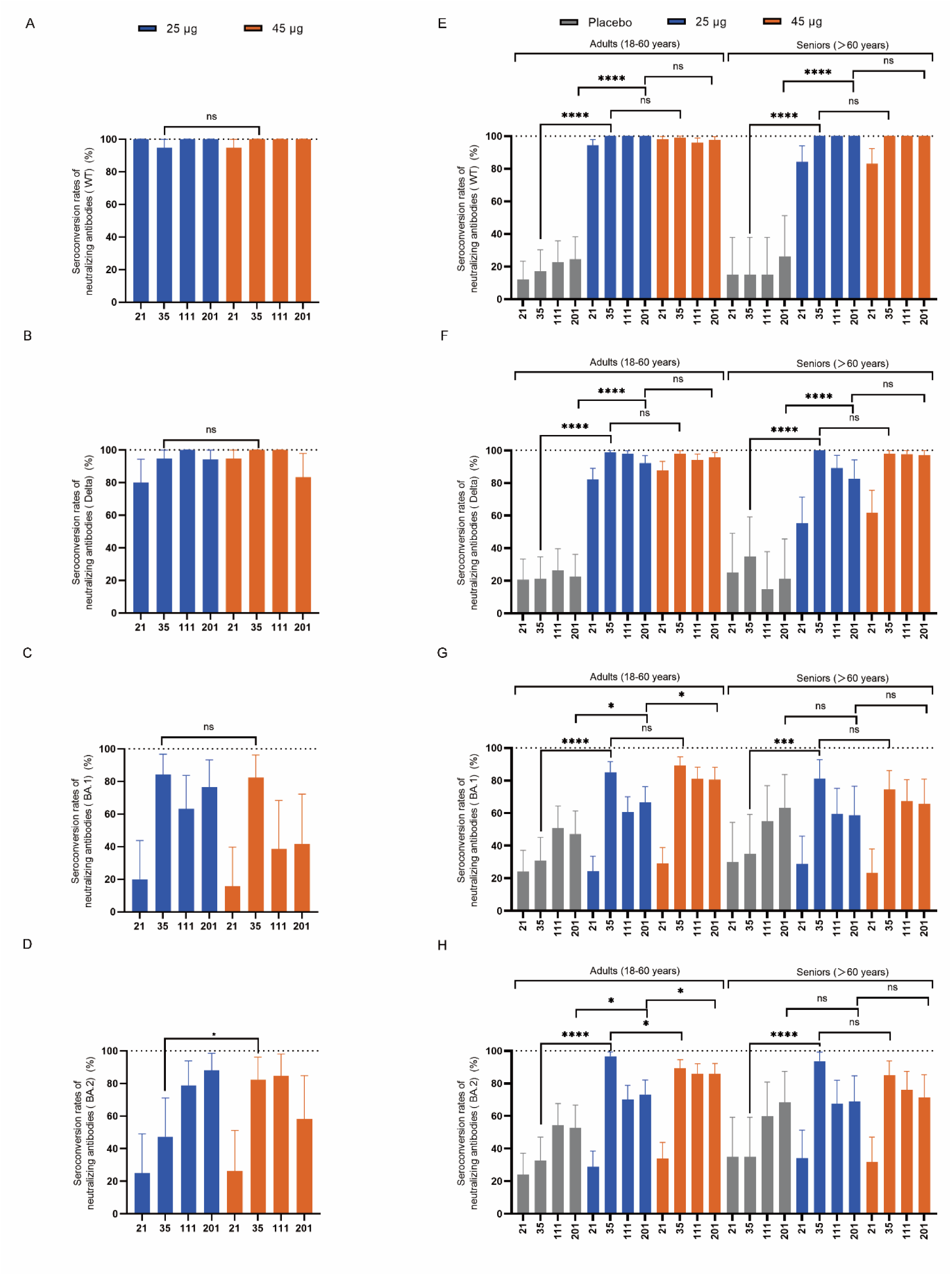
Seroconversion rate of SW-BIC-213 induced pseudo-virus neutralizing antibodies in phase 1 and phase 2 trials. A. The seroconversion rate of SARS-CoV-2 prototype strain pseudo-virus neutralizing antibody at the indicated time points (day) in phase 1 trial. B. The seroconversion rate of SARS-CoV-2 delta strain pseudo-virus neutralizing antibody at the indicated time points (day) in phase 1 trial. C. The seroconversion rate of SARS-CoV-2 omicron BA.1 strain pseudo-virus neutralizing antibody at the indicated time points (day) in phase 1 trial. D. The seroconversion rate of SARS-CoV-2 omicron BA.2 strain pseudo-virus neutralizing antibody at the indicated time points (day) in phase 1 trial. E. The seroconversion rate of SARS-CoV-2 prototype strain pseudo-virus neutralizing antibody at the indicated time points (day) in phase 2 trial. F. The seroconversion rate of SARS-CoV-2 delta strain pseudo-virus neutralizing antibody at the indicated time points (day) in phase 2 trial. G. The seroconversion rate of SARS-CoV-2 omicron BA.1 strain pseudo-virus neutralizing antibody at the indicated time points (day) in phase 2 trial. H. The seroconversion rate of SARS-CoV-2 omicron BA.2 strain pseudo-virus neutralizing antibody at the indicated time points (day) in phase 2 trial. Seroconversion is defined as titers above or equal 10 in baseline-seronegative subjects or at least 4-fold rise in baseline-seropositive subjects. Chi-square test was used to analyze the difference of seroconversion rate of the indicated group. The results of compared analysis among the three groups were expressed as no significant difference ns (P>0.5), * (p≤0.05), ** (p≤0.01), *** (p ≤0.001), **** (p≤0.0001).

